# Finger stick blood test to assess post vaccination SARS-CoV-2 neutralizing antibody response against variants

**DOI:** 10.1101/2021.11.11.21266241

**Authors:** Sing Mei Lim, Hoi Lok Cheng, Huan Jia, Patthara Kongsuphol, Bhuvaneshwari D/O Shunmuganathan, Ming Wei Chen, Say Yong Ng, Xiaohong Gao, Shuvan Prashant Turaga, Sascha P. Heussler, Joyti Somani, Sharmila Sengupta, Dousabel MY Tay, Megan E. McBee, Barnaby E. Young, Paul A. MacAry, Hadley D. Sikes, Peter R. Preiser

## Abstract

There is clinical need for a quantifiable point-of-care (PoC) SARS-CoV-2 neutralizing antibody (nAb) test that is adaptable with the pandemic’s changing landscape. Here, we present a rapid and semi-quantitative nAb test that uses finger stick or venous blood to assess the nAb response of vaccinated population against wild-type, alpha, beta, gamma, and delta variant receptor binding domains. It captures a clinically relevant range of nAb levels, and effectively differentiates pre-vaccination, post 1^st^ dose and post 2^nd^ dose vaccination samples within 10 minutes. The data observed against alpha, beta, gamma, and delta variants agrees with published results evaluated in established serology tests. Finally, our test revealed a substantial reduction in nAb level for beta, gamma, and delta variants between early BNT162b2 vaccination group (within 3 months) and later vaccination group (post 3 months). This test is highly suited for PoC settings and provides an insightful nAb response in a post-vaccinated population.

## 1 INTRODUCTION

Highly transmissible SARS-CoV-2 variants such as B.1.1.7 (alpha) and B.1.617.2 (delta) have emerged and displaced the ‘wildtype’ virus and other variants within countries with high vaccination rates. With 38.9 % of global population now vaccinated (as of 5th Nov 2021, live update from https://ourworldindata.org/covid-vaccinations), reports of breakthrough infections among vaccinees indicate the potential need for future vaccine boosters, particularly in vulnerable populations ^1–3^. A rapid, easy to use Point-of-Care (PoC) test that measures the level of immune protection against SARS-CoV-2 in both recovered as well as vaccinated individuals over time would be an important tool in guiding public health policy. Currently, standard viral neutralization test (VNT) and pseudovirus neutralization test (pVNT) have played critical roles in evaluating protective immunity, however their use is limited due to the need for BSL2 or BSL3 laboratory facilities, extended experimental time and relevant expertise. Moreover, the reproducibility varies depending on cell type, virus/pseudovirus generation, experimental protocol, and detection method ^4,5^. While ELISA-based surrogate neutralization test can provide reliable information on immune protection, it requires skilled operators and dedicated facilities that are difficult to integrate into PoC testing ^6,7^. PoC lateral flow tests are currently limited, as they either detect total immunoglobulin level which is not a reliable indicator for immune protection or only provide qualitative assessment ^8,9^. The availability of a quick and accurate PoC nAb test to track vaccination induced immune responses especially against variants at both the population as well as individual level would be a valuable tool in enabling public health authorities to manage breakthrough infections and to develop an effective booster vaccination strategy for more vulnerable individuals.

We previously developed a rapid paper-based SARS-CoV-2 neutralization assay known as cellulose pulled-down virus neutralization test (cpVNT) that detects SARS-CoV-2 neutralizing antibody (nAb) in plasma or serum within 10 minutes ^10^. The principle of cpVNT is based on the complex formation between the receptor binding domain (RBD) of the SARS-CoV-2 and the angiotensin converting enzyme II receptor (ACE2) of the host cell. The RBD is fused with cellulose binding domain (RBD-CBD) to enable capture by cellulose paper while ACE2 is conjugated with reporting molecules for signal generation. The presence of nAb in the sample disrupts RBD-CBD/ACE2 complex formation leading to a reduction in the overall signal detected. To develop a PoC nAb detection test, we improved our cpVNT assay enabling it to be used directly on whole venous or capillary blood including finger stick blood samples. This bypasses the requirement for extensive sample processing or a phlebotomist. Comparison of this modified cpVNT test with established pVNT as well as an ELISA-based assay showed high degree of concordance. Importantly, the modified cpVNT test can be easily adapted for the rapid evaluation of nAb responses to SARS-CoV2 variants among vaccinated population, providing critical insights into changes in nAb responses to vaccine types, variant mutations, and time post vaccination.

## 2 RESULTS

### 2.1 Rapid detection of SARS-CoV-2 neutralization antibody in blood using modified cpVNT

To adapt the previous cpVNT for the analysis of whole blood PoC diagnostic samples it was important to change the enzyme/substrate-based reporter system of HRP/TMB as well as the overall assay workflow. The two key modifications introduced are (i) the change to a fluorescent reporter molecule and (ii) sequential incubation steps (Fig. 1A, S1A). For this, we selected Alexa Fluor® 594 as the reporter in consideration of its high quantum yield, excellent photostability, and minimal interference with blood. In addition, to improve the test performance and minimize the non-specific background from whole blood sample, we altered the cpVNT workflow to a two-step incubation, as compared to the one-step incubation reported previously. First, 20 µL of blood sample was mixed with 20 µL of 10 nM RBD-CBD for 3 minutes before adding 40 µL of 5 nM fluorescence labelled human ACE2 (ACE2-AF594) and incubating for additional 5 minutes at ambient temperature. Equal amount of the final 80 µL mixture was then applied to the cassette’s test and control spot respectively followed by one washing step with 40 µL of PBS for each spot (Fig. 1A & B). The additional 3 minutes incubation step introduced in this study allowed nAb in the blood sample to effectively interact with RBD-CBD prior to exposure to ACE2 (Fig. S1 B & C).

**FIGURE 1.**
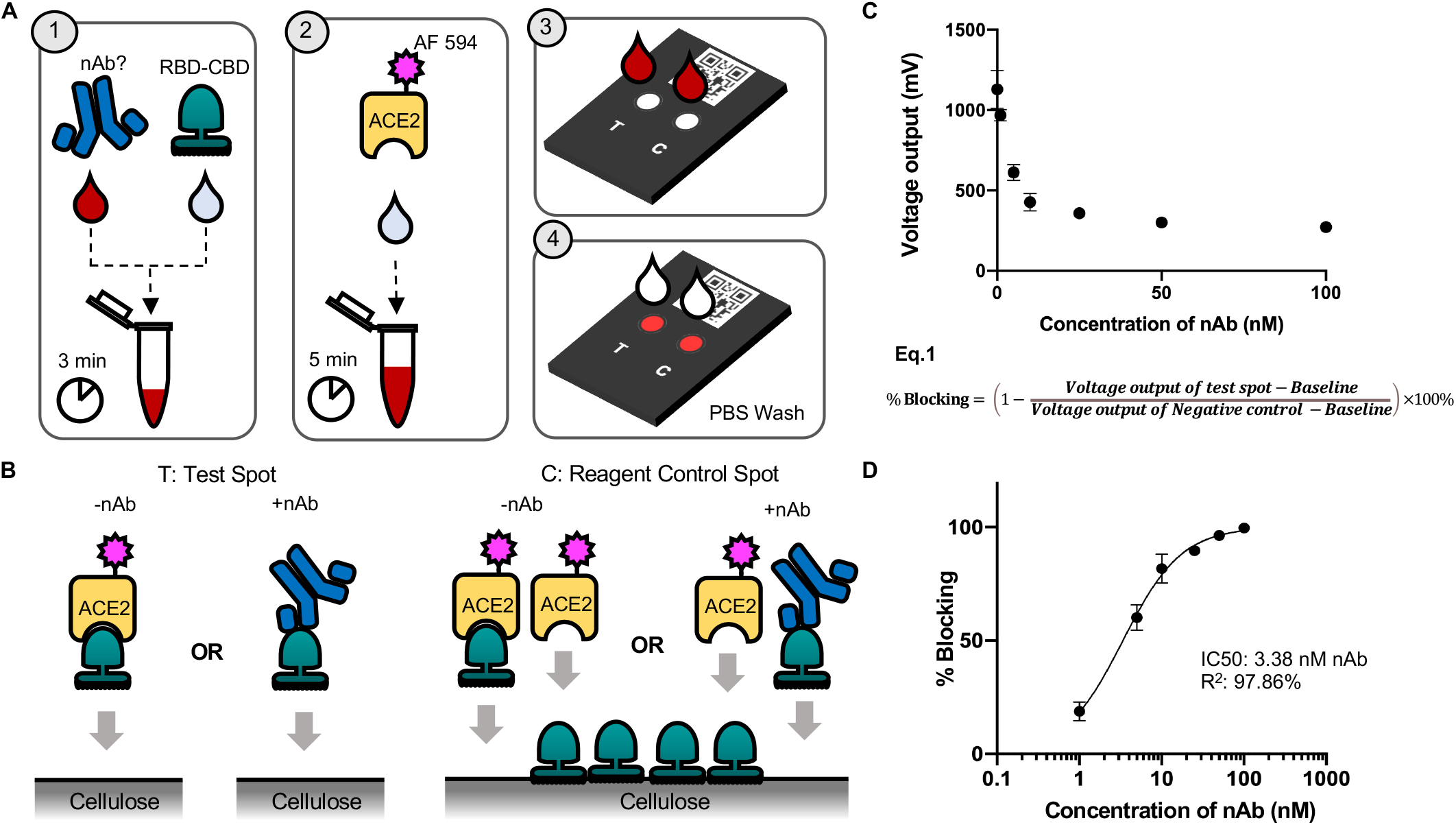
Schematic of cpVNT workflow and results obtained with contrived whole-blood sample. **A**. Graphical representation of the modified cpVNT workflow before detection in fluorescent reader. **B**. Cartoon depicting of possible molecular events occur in samples with and without nAb on the test spot and reagent control spot. **C**. Measurement of fluorescence intensity from pre vaccination whole blood samples titrated with 1, 5, 10, 25, 50, 100 nM SARS-CoV-2 monoclonal neutralizing antibody (nAb) **D**. The percent blocking calculated from Equation 1 with nAb concentration presented in log scale.

Since the presence of nAb is inversely related to the fluorescence intensity, a control reaction is necessary to verify the reagent functionality, as the loss of signal should only be due to the presence of nAb. Therefore, we pre-immobilized the control spot with 5 µL of 5 µM of RBD-CBD on the cellulose paper to capture ACE2-AF594 free from RBD-CBD/ACE2-AF594 complex and produce high level of fluorescent signal regardless of the level of nAb present in the blood (Fig. 1A, S1 D). A portable fluorescent reader, Atto Testbed produced by Attonics Systems Pte. Ltd., Singapore is customized to allow the detection of fluorescence signal under a PoC setting. The reader excites the fluorophores using LED light. The emitted fluorescent intensity is then detected using a silicon avalanche photodiode and reported as a voltage change in mV unit. This voltage output (mV) can be converted to percentage of blocking according to Equation 1:

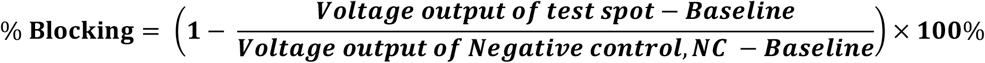

To evaluate this new test format, we made a series of contrived blood samples by spiking 1, 5, 10, 25, 50 and 100 nM of mouse monoclonal SARS-CoV-2 nAb into a blood prepared with pre-SARS-CoV-2 pandemic plasma and washed red blood cells. The assay demonstrated an IC_50_ of 3.38 nM nAb using blood as sample matrix (Fig. 1C & D).

### 2.2 Evaluation of post vaccination nAb responses using modified cpVNT

Modified cpVNT relies on the reduction of fluorescent signal in response to neutralizing antibodies competing with labelled ACE2. Two important fluorescent signals contribute to the determination of the signal dynamic range and nAb result interpretation, (i) the maximum fluorescence intensity obtained from negative control (NC) samples with no nAb (pre-vaccination, Pre-Vac samples); this value was used to set a reference point for calculation of blocking percentage in the presence of nAb and (ii) non-specific background fluorescence observed from the test when RBD-CBD was absent from the reaction; this value was used to draw a baseline between specific and non-specific signals (see Equation 1). The NC value was defined by the median of fluorescence intensity measured from 60 Pre-Vac blood samples in triplicates (Fig S2A), while the baseline value was the median of triplicate reads from 31 blood samples regardless of vaccination status when RBD-CBD was absent (Fig S2B). With this approach, the NC value using two independent batches of ACE2-AF594 resulted in a median fluorescence intensity of 1141 mV (Table S1 and Fig S2A) while the baseline signal had a median intensity value of 230 mV (Fig S2B). To calculate the percentage of blocking based on the nAb levels of individual, we then applied the NC as well as baseline value to formulate Equation 1.

Once NC and the baseline were established, the nAb levels that block RBD and ACE2 interaction were measured in 170 blood samples using modified cpVNT at different stages of vaccination: pre-vaccination (Pre-Vac), 1-2 weeks post 1^st^ dose (P1 1-2W), 3-6 weeks post 1^st^ dose (P1 3-6W) and 3-16 weeks post 2^nd^ dose (P2). The Pre-Vac (n=36) group’s percent blocking was measured with a median of 1.96%, this number increased to 14.3% in P1 1-2W group (n = 10) and P1 3-4W group at 36.5% (n = 50). Lastly, we observed 89.1% for P2 samples (n=74) (Fig. 2A). A similar trend was observed when grouping the samples into individuals vaccinated with either BNT162b2 (Fig 2B) or mRNA-1273 (Fig 2C). The negative value of percent blocking observed in the Pre-Vac and P1 1-2W was likely due to viscosity variation among blood samples that could interfere with the binding kinetics of RBD-CBD with cellulose paper in the assay. Overall, the data showed a significant difference between Pre-Vac and P1 3-6W samples across the two different types of vaccines, i.e. BNT162b2 (p<0.05) and mRNA-1273 (p<0.001) (Fig 2B & C). There was also a significant difference (p<0.05) in median percent blocking for P1 3-6W group when compared between BNT162b2 recipients (23.4%, n=30) and mRNA-1273 recipients (51.2%, n=20) (Fig 2D). However, in P2 samples, both vaccines show comparable median percent blocking at 84.5% for BNT162b2 and 90.9% for mRNA-1273 recipients (Fig 2B & C). Mapping of the nAb response in 22 individuals who received either BNT162b2 or mRNA-1273 showed differential responses at P1 phase (Fig 2E). Both vaccines induced a heterogeneous though elevated response in all individuals tested as early as week 2 post first dose of vaccine.

**FIGURE 2.**
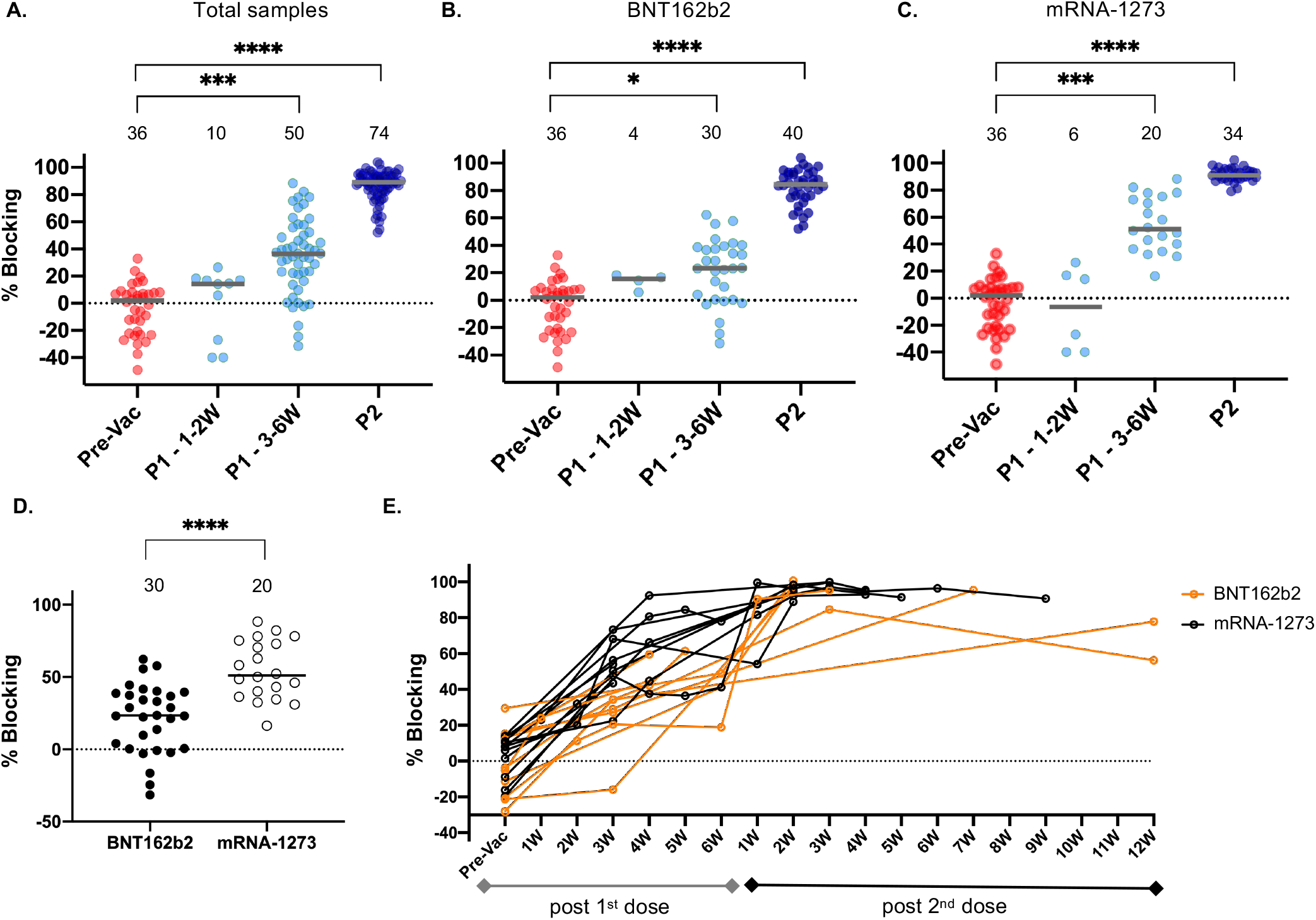
Evaluation of nAb response among pre-vaccination and post-vaccinated individuals using modified cpVNT. **A**. The percent blocking measured from 170 whole blood samples of pre vaccination, Pre-Vac (n=36), post 1^st^ dose (1-2W n=10, 3-6W n=50) and post 2^nd^ dose (n=74). The grey line is the median from each group while each dot represents the mean from three independent experiments. **B**. The nAb percent blocking measured in individuals opt for BNT162b2 (n=4 from P1 - 1-2W, n=30 from P1 - 3-6W, n=40 from P2) or **C**. mRNA-1273 (P1 - 1-2W n=6, P1 - 3-6W n=20, P2 n=34). Kruskal-Wallis test with Dunn’s multiple comparison was performed between each vaccination status. **D**. Comparison of nAb percent blocking at P1 3-6W BNT162b2 and P1 3-6W mRNA-1273. Two tailed Mann-Whitney test was performed between the two vaccine brands. The significance values * *P* < 0.05, ** *P* < 0.01, \*\*\**P* < 0.001, \*\*\*\**P* < 0.0001. Total samples: Pre-Vac versus P1 – 3-6W, *P* < 0.001; Pre-Vac versus P2, *P* < 0.0001. BNT162b2: Pre-Vac versus P1 – 3-6W, *P* < 0.05; Pre-Vac versus P2, *P* < 0.0001. mRNA-1273: Pre-Vac versus P1 – 3-6W, *P* < 0.001; Pre-Vac versus P2, *P* < 0.0001. **E**. Twenty-two individual samples percent blocking mapped over pre and post vaccination period comparing between two types of vaccines, BNT162b2 n=12, mRNA 1273 n=10. The window period between first and second dose of vaccination ranged from 4 to 6 weeks depending on individual’s choice.

### 2.3 Modified cpVNT can detect wide range of nAb activities comparable to sVNT and pVNT

We used the WHO International Standard for anti-SARS-CoV-2 immunoglobulin (20/136) and Reference Panel (20/268) to assess the detection range of modified cpVNT and to better interpret the clinical data we have measured ^11^. The International Standard comprising of plasma sample with assigned 1000 IU/mL nAb activity resulted in 96.7% blocking in our modified cpVNT (Fig 3A). This percent blocking corresponded to the value observed from subjects in the P2 vaccination group (Fig. 2A). The Mid-titre and Low-titre plasma from WHO Reference Panel with 210 IU/mL and 44 IU/mL nAb activity respectively were measured with 66.7% blocking (Mid) and 34.9% blocking (Low) in the modified cpVNT (Fig 3A). This showed that the test can produce a dose dependent response that captures the clinical range of nAb activity in plasma in under 10 minutes assay time. Since plasma represents approximately 55% of whole blood, the percent blocking test results in plasma samples was expected to be higher than that of whole-blood due to the lack of erythrocytes. To correlate the percent blocking in the WHO standard and reference panel plasma to corresponding whole blood, we analyzed 30 matching samples of blood and plasma in the modified cpVNT. We found that the percent blocking in blood samples is approximately 0.87 times of that in plasma samples assuming the relationship between the two sample types are linear (Fig 3B). We observed that the overall median percent blocking in Pre-Vac samples was found below 30% blocking in modified cpVNT using blood as matrix (Fig 2A). It corresponds to 44 IU/mL neutralizing antibody activity which is close to the estimated protective neutralization against SARS-CoV-2 of approximately of 54 IU/mL by Khoury et al.’s predictive model ^12^ (Fig 3B). Hence, 30% blocking which correlates to 44 IU/mL was set as the cut-off value for the modified cpVNT to compare its performance with other neutralization tests (Fig 3B). Our test showed 81.5% sensitivity (CI 61.9-93.7%) and 100% specificity (CI 81.5-100%) when compared with the commercially available sVNT Genscript cPass™ (Fig 3C). Meanwhile, as compared to the lab based pVNT test, the modified cpVNT showed 100 % sensitivity (CI 47.8 - 99.9%) and 66.7% specificity (CI: 38.4 - 88.2%) (Fig 3D). For reference, the WHO plasma of nAb activity at 1000 IU/mL, 210 IU/mL and 44 IU/mL when performed with sVNT cPass yielded 94 %, 78% and 19% inhibition respectively (Table S3). The lower specificity and sensitivity relative to ELISA and pVNT can be attributed to the difference in sample type (whole blood vs plasma/serum) and different assay procedures.

**FIGURE 3.**
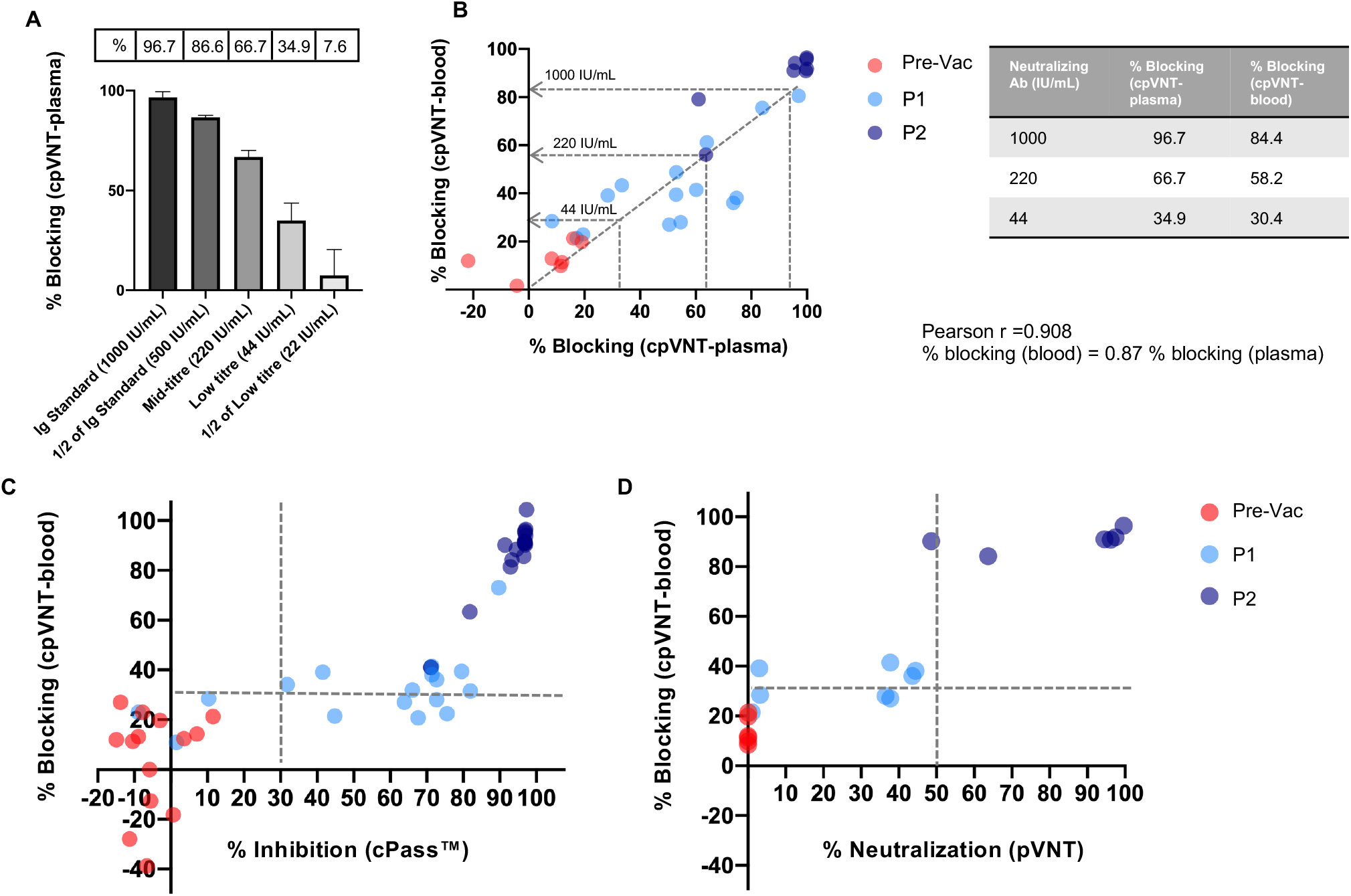
Comparison of modified cpVNT with international standards and established serology tests. **A**. The performance of the First WHO International standard Anti-SARS-CoV-2 Immunoglobulin (20/136), Reference Panel for anti-SARS-CoV-2 Mid-tire and Low-titre plasma using modified cpVNT. **B**. The correlation of percent blocking measured from 30 matching plasma and blood samples at pre vaccination (Pre-Vac), post 1^st^ dose (P1) and post 2^nd^ dose (P2) phase using modified cpVNT gave Pearson r, 0.908. The percent blocking for cpVNT with blood samples that correspond to 1000 IU/mL, 220 IU/mL and 44 IU/mL are determined by assuming a linear correlation between the two sample types (see accompanying table). **C**. Comparison of percent blocking measured in the modified cpVNT with percent inhibition of sVNT (cPass™) in 45 matching Pre-Vac, P1 and P2 venous blood and plasma samples. The sensitivity was calculated as 81.5% (CI: 61.9-93.7%), and specificity is 100% (CI: 81.5-100%) when both cpVNT and sVNT’s (cPass™) threshold were set at 30% blocking. **D**. Comparison between cpVNT and pseudovirus neutralization test (pVNT) with 20 individuals’ sample. The pVNT was performed with plasma in 1:80 dilution. The sensitivity is 100% (CI: 47.8-99.9%) and specificity is 66.7% (CI: 38.4-88.2%) with 30% blocking as a threshold for cpVNT while 50% neutralization for pVNT. All experiments were performed in triplicates.

To ensure that the modified cpVNT is suitable for PoC setting with finger stick blood, we assessed the correlation of nAb detected in venous vs finger-stick blood sample matrix. A total of 46 matched pairs from Pre-Vac, P1 and P2 samples show a high linear correlation between the two blood sample types, with a Pearson r value of 0.9758 (p value < 0.001) and an R^2^ of 0.9523 (Fig S2C). This demonstrates the suitability of our modified cpVNT for PoC deployment, as only 20 µL of finger-stick blood samples is necessary to measure the nAb response in 10 minutes. Moreover, the outcomes are comparable to established lab-based neutralization tests.

### 2.4 Assessment of post-vaccinated nAb percent blocking against SARS-CoV-2 variant RBDs

Given the emergence of several SARS-CoV-2 variants and the accompanying uncertainty of the effectiveness of vaccine-induced nAbs against them, there has been burgeoning interest in evaluating nAb responses to variant RBDs. In light of this, we first recombinantly produced 4 variants of concern (VOCs): alpha B.1.1.7, beta B.1.351, gamma P.1, delta B.1.167.2 and 5 variants of interest (VOIs): kappa B.1.167.1, epsilon B.1.427/B.1.429, delta plus AY.1, eta B.1.525, lambda C.37 fused with CBD and evaluate their binding activity with ACE2 using Biolayer-Interferometry (BLI) (Fig. S3 & S4A). These variants contain mutations in the RBD region, which may reduce the binding affinities of antibodies generated against the wildtype protein and/or increase ACE2 receptor binding ^13^. We found that the binding affinity of alpha, beta, gamma, and delta are higher than that of wildtype (WT), especially gamma that showed a 3 -fold increase (4.3 nM) in binding affinity comparing to wild-type (12.7 nM) consistent with previous report ^13,14^ (Table 1). Furthermore, our result supports published data that the N501Y mutation in the alpha, and gamma variants of RBD contributes to the slow off-rate of the complex ^15^ (Table 1). Meanwhile T478K appears to promote fast complex formation based on comparison among delta, kappa, epsilon, and a delta plus variant that shared the L452R mutation (Table 1). We engineered a hypothetical RBD variant containing N501Y, T478K mutation and annotated it as ‘AD’ (alpha-delta) variant that is speculated to have fast on-rate and slow off-rate with ACE2. This hypothetical variant confirmed our hypothesis where it binds ACE2 with the highest binding affinity (KD of 3 nM) among the 10 variants (Table 1). Next, we assess the activity of these RBD-CBD variants on the modified cpVNT with Pre-Vac blood. Although variants with high affinity to ACE2 showed increased fluorescence intensity than WT RBD-CBD in the modified cpVNT assay at the same reagent concentration, ie. alpha (1.8-fold), beta (1.6-fold), gamma (2.1-fold), delta (1.3-fold), the correlation is not direct. As we observed variant RBD-CBDs epsilon and lambda still generate comparable signal as WT despite the lower ACE2 binding affinity, while AD variant showed merely 1.6-fold increase in signal despite binding ACE2 strongly (Fig. S4A). Besides the binding kinetics, the capture rate of RBD-CBD on the cellulose paper and possible avidity of the different RBD-variants on ACE2 could contribute to the effect.

**TABLE 1:**
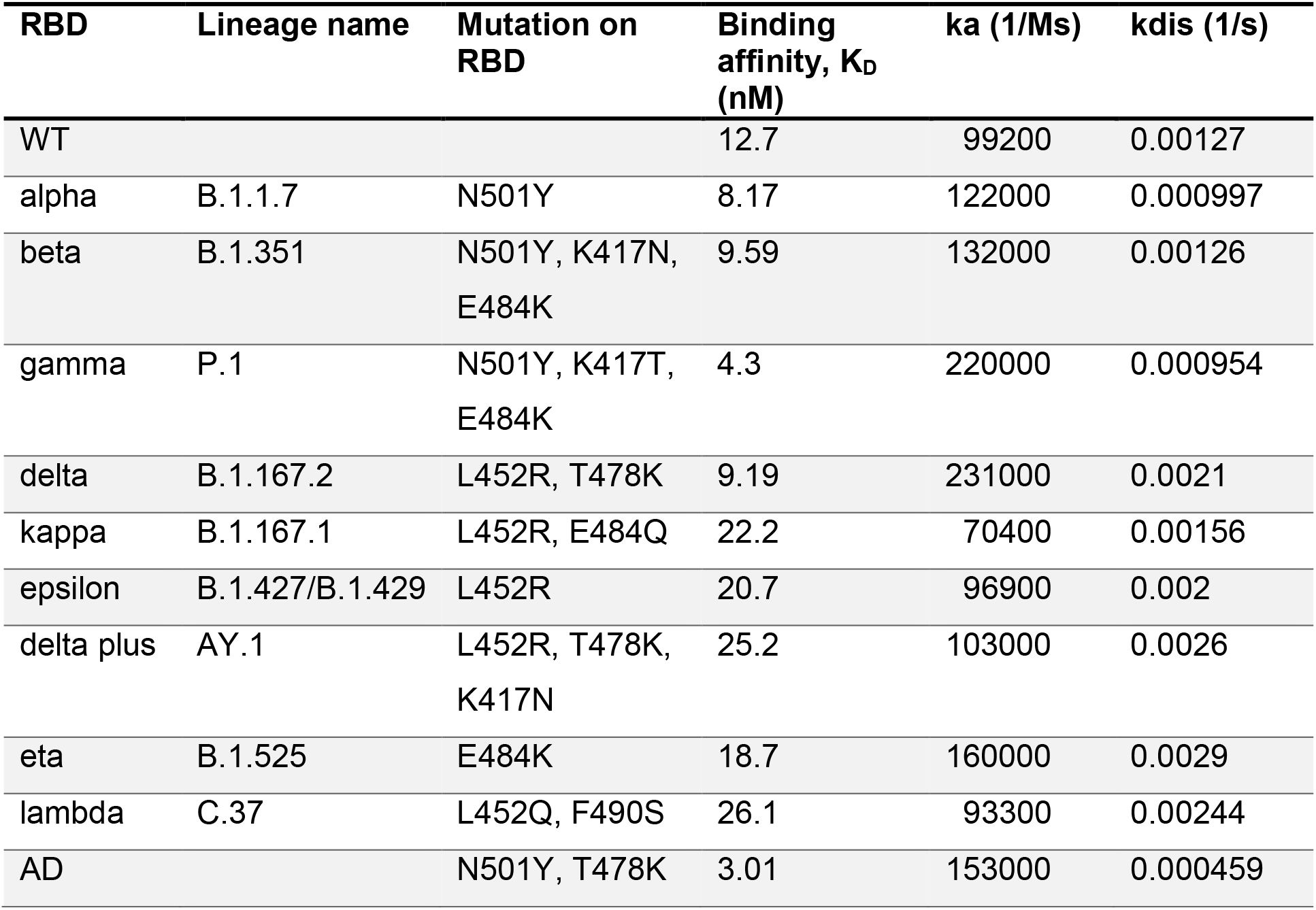
Binding kinetics of wildtype and RBD-CBD variant with biotinylated ACE2.

We then tested the 4 VOCs: RBD-CBD alpha, beta, gamma, and delta with 33 blood samples from participants within 3 months of completing vaccination. There were considerable variations in the nAb responses to the different variants. The nAb percent blocking against beta and gamma variants being reduced significantly to 72.4 % and 70.1%, while the percent blocking reduced only minimally to 87.2% and 91.9% for alpha and delta respectively as compared to WT (95.6%) (Fig 4A). These were in line with previous reports using pVNT and VNT, whereby neutralization of beta and gamma variants had considerable reduction for both mRNA vaccines ^16–18^. About 91.8% nAb blocking was observed against the engineered AD variant even though the RBD-CBD variant binds strongly to ACE2, suggesting that vaccine induced nAb can outcompete stronger interaction (Fig S4B). While this data indicated a heterogenous response it was important to evaluate whether our test was able to stratify response in relation to the different vaccines used. The median percent blocking for BNT162b2 recipients against alpha was 78% (p< 0.01) and delta was 89.2% (p = n.s.) as compared to WT (94.8%). (Fig 4B). The most substantial reductions of nAb response were observed with beta and gamma variants reaching 55.7% and 49.6% blocking respectively among BNT162b2 recipients (Fig 4B). In the cohort of mRNA-1273 recipients, we observed reduction to 87.5% with beta variant (p<0.0001) and 80.5% with gamma variant (p <0.0001).

**FIGURE 4.**
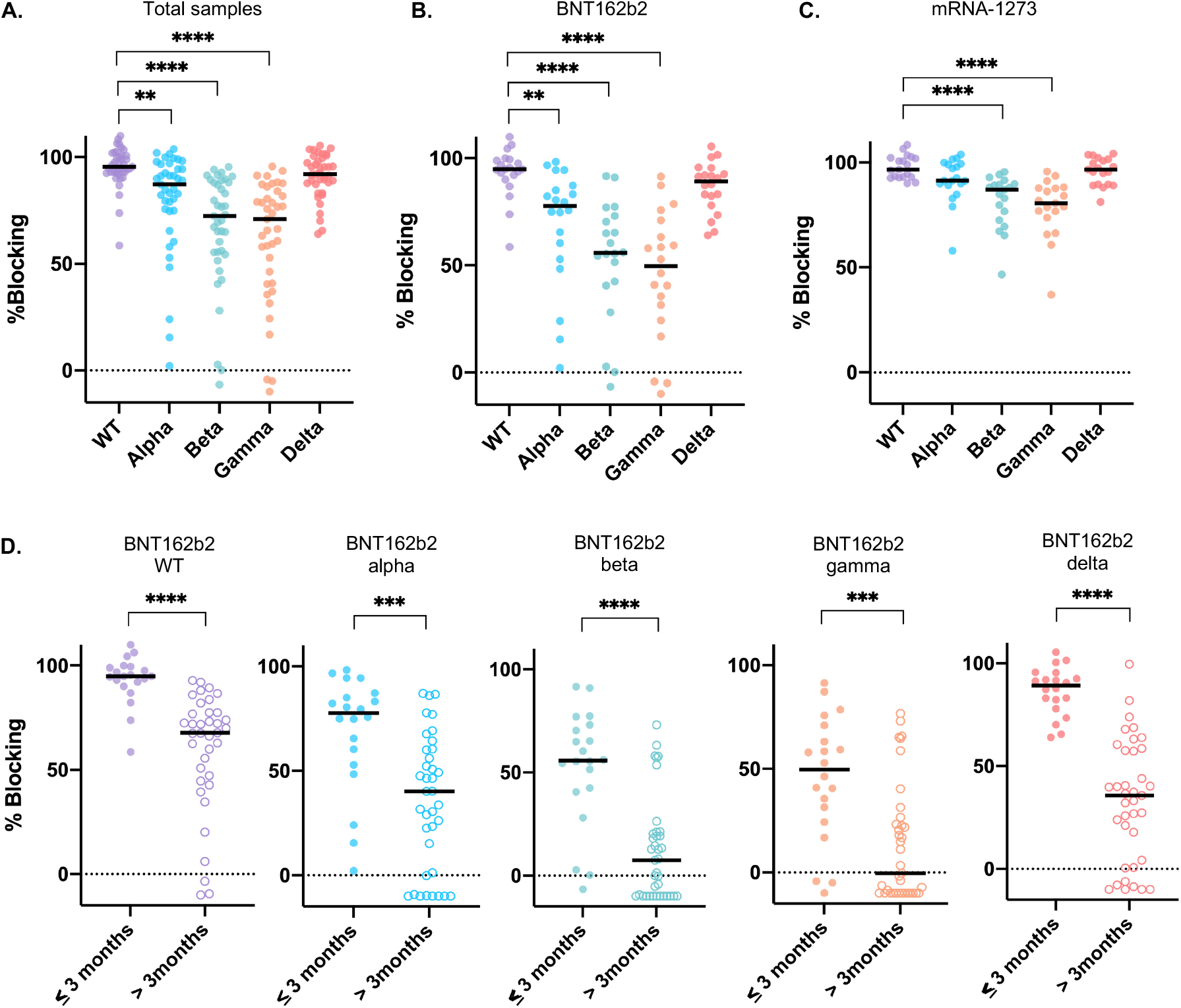
Assessment of nAb response to variants of concern depending on vaccine brand or time post vaccination with our modified cpVNT. **A**. Whole blood samples (n=39) from participants who completed vaccination within three months were tested with wildtype (WT) RBD-CBD and variant RBD-CBD. Friedman test with multiple comparison was performed comparing the variants against WT, \**P* < 0.05, \*\**P* < 0.01, \*\*\**P* < 0.001, \*\*\*\**P* < 0.0001. WT versus alpha, *P* < 0.01. ; WT versus beta, *P* < 0.0001 ; WT versus gamma, *P* < 0.0001; WT versus delta, n.s. **B**. The percent blocking of nAb measured from 20 P2 samples of BNT162b2 recipients (≤ 3 months) when tested with RBD-CBD variants **C**. Similarly, percent nAb blocking of 19 P2 samples (≤ 3 months) from mRNA-1273 recipients tested with the RBD-CBD variants. **D**. The nAb response in BNT16b2 recipients against WT and RBD-CBD variants when comparing two groups: within three months (n=20) and more than three months (n=37) after complete vaccination. Two-sided Mann Whitney test was performed between the two groups for each variant, WT *P* < 0.0001, alpha *P* < 0.001, beta *P* < 0.0001, gamma *P* < 0.001, delta *P* < 0.0001.

Next, we also examined and compared the percent blocking of nAb in whole-blood samples from participants within three months or greater than three months after completion of vaccination against WT and 4 VOCs RBD-CBD. Only samples from BNT162b2 recipients were available to us for the greater than three months cohort as it was the first vaccine rolled-out in the Singapore national vaccination program. There was a modest drop of nAb percent blocking from 96% to 68% (28%, p<0.0001) observed in WT RBD-CBD and 77.6% to 40.2% (37.4%, p<0.001) in alpha RBD-CBD between the two groups of samples (Fig 4D). Meanwhile a more substantial reduction was seen in beta (47.8%, p <0.0001), gamma (49.5%, p <0.001) and delta variant (53.5%, p <0.0001) respectively (Fig 4D). It is interesting to observe the stark decline of nAb blocking for delta variant in the post 3 months cohort especially when no significant difference of nAb response was detected from WT for samples vaccinated within 3 months (Fig 4A). The modified cpVNT results agree with recent findings where BNT162b2 vaccine induced protection wanes within 4-5 months especially against the delta variant despite earlier findings suggest effective neutralization ^19,20^. This demonstrates that this test can be effectively adapted in the event of future VOC emergence to quickly assess vaccinees’ responses and even to identify vulnerable individuals for booster shots to prevent breakthrough infections.

## 3 DISCUSSION

The rapid modified cpVNT can improve our understanding of the relationship between nAb response and RDB/ACE2 interaction, especially in response to emerging and predictive mutants. Given the complex innate and cell mediated immune response against infection and immune-protection development, factors like synergistic mutations and epitope remodelling to prevent nAb recognition are key to a variant’s immune escape characteristics ^21^. We tested the influence of RBD/ACE2 interaction on nAb blocking with the hypothetical AD variant that carries N501Y T478K mutation. It was found unable to evade vaccine induced nAb inhibition where it shows 91.8% nAb blocking in the modified cpVNT similar to WT RBD-CBD despite its high affinity to ACE2 (Fig S4B). Since a single T478K mutation did not present compromising effect on the binding of potent neutralizing mAbs previously ^22^, we observed that the additional N501Y mutation in AD variant does not affect nAb binding within the modified cpVNT’s reaction time (Fig S4B). In contrast, the beta variant despite showing modest increase in affinity towards ACE2 (K_D_ 9.6 nM) than WT (K_D_ 12.7 nM), exhibited significantly lower nAb percent blocking than WT (Fig. 4A). As the K417 and E484 sites are known to escape both class 1 and class 2 anti-RBD antibodies ^23^, the combined effect of RBD/ACE2 binding and poor nAb recognition generate more pronounced immune escape response. These examples indicate that the modified cpVNT can be used to systematically assess the RBD mutations and improve our understanding of its underlying molecular mechanism versus nAb response.

With the emergence of highly transmissible SARS-CoV-2 variants, the durability and persistence of vaccine effectiveness is of major concern. Although nAb response strongly correlates with immune protection ^12^, cellular immunity is essential in providing sustained immune protection upon exposure, particularly against severe illness. Therefore, both humoral and cellular immune response are required for a complete assessment of SARS-CoV-2 immunity. While standardized methods for rapid assessment of cellular immunity responses are underway ^24^, nAb level measurement remains a reliable indicator for immune-protection at PoC level and deems to be critical at this point. It has been estimated that 90% of convalescent plasma/sera’s neutralizing activity targets the immunodominant RBD ^25–27^, hence the current modified cpVNT format that measures the nAb response to RBD-associated mutations represents a good proxy for assessing individual’s immune protection. The standardized percent blocking provided by the customized reader, permits consistent results interpretation as opposed to colorimetric scoring. Besides, as demonstrated here, the test only requires a simple change in one reagent while retaining the test format, instrumentation, and capability to evaluate nAb responses to a new variant. This feature is quintessential for keeping up with the rapidly evolving virus, for example the new mu variant in Colombia that was reported to escape vaccine induced immunity^28^.

Our data also shows the strength of the modified cpVNT as a PoC test to provide insights on the deteriorating vaccine efficacy observed globally against the delta variant and the climbing breakthrough infections among vaccinated population. The significant decline of nAb response against the delta variant observed among post 3 months’s BNT162b2 vaccinees in our study provides a possible reason for the increased breakthrough infections observed globally. The report that BNT162b2 recipients who completed their vaccination between Jan-April in Israel had an increased risk of breakthrough infections with delta variant is in line with our interpretation ^29^. Thus, our test’s ability to detect variant specific nAb waning effects among a vaccinated population provides an extremely valuable tool to pre-emptively test nAb responses against emerging variants and through this inform booster planning and public health management.

## 4 MATERIALS AND METHODS

### 4.1 Study participants

Heathy adults age between 21-65 years old scheduled for Singapore national vaccination program were enrolled to the study in compliance with all relevant ethical regulations and was approved by Institutional Review Board of Nanyang Technological University (IRB-2021-04-020). All participants provided informed consent before participation under voluntary basis and reported with no prior SARS-CoV-2 infection at point of recruitment. The venous blood collection was performed by certified phlebotomists while finger-prick blood was collected using Haim® Winnoz blood collection device or manual collection. Pre-SARS-CoV-2 plasma samples were collected under IRB 003/2010, IRB 11/08/03, IRB 13/09/01 and IRB-2016-01-045 stored in -80°C. Whole-blood samples from healthy volunteers vaccinated more than 3 months was provided by National Centre of Infectious Diseases (NCID) under DSRB 2012/00917. There were no breakthrough infections reported from these samples.

### 4.2 Blood sample processing and storage

Blood samples were kept at 4°C for delivery, venous blood storage in heparin tubes (BD Vacutainer® #367874) while finger stick blood were stored in either heparin (Xinle Medical MP0540) or EDTA (Xinle Medical MP0581) microtainer tubes. A portion of the sample volume was separated into plasma content by centrifugation at 4000 g for 5 minutes in 4°C. Plasma were stored in -20°C. Both WHO International Standard (20/136) and Reference Panel for anti-SARS-CoV-2 immunoglobulin (20/268) plasma were purchased from National Institute for Biological Standards and Control (NIBSC, United Kingdom) and were stored in -20 °C upon receipt.

### 4.3 Protein production and purification

The expression and purification of soluble extracellular fragment of human ACE2 (residues 19–615; GenBank: AB046569.1) and wildtype (WT) SARS-CoV2-Spike (EMBL: QHD43416.1 with silent mutations c.A1452>G and c.T1470>C) RBD fused to CBD followed the same protocol as described in Kongsuphol et al. ^10^. Similarly, alpha c.A1501>T (p.N501Y), beta c.A1501>T, c.G1251>C, c.G1450>A (p.N501Y K417N E484K), gamma c.A1501>T, c.A1250>C, c.G1450>A (p.N501Y K417T E484K), delta c.T1355>G, c.C1433>A (p.L452R T478K), kappa c.T1355>G, c.G1450>C (p.L452R E484Q), epsilon c.T1355>G (p.L452R), delta plus c.T1355>G, c.C1433>A, c.G1450>A (p.L452R T478K K417N), eta c.G1450>A (p.E484K), lambda c.T1355>A c. T1469>C (p.L452Q, F490S) and AD c.A1501>T, c.C1433>A (p.N501Y T478K) RBD-CBD variants were expressed in Expi293F cells (Thermo Fisher Scientific, A1435101) according to the supplier’s protocol. The purification protocol followed that of WT RBD-CBD. In brief, the proteins were subjected to affinity chromatography with Ni-NTA cartridges (Qiagen, 1046323) and size exclusion chromatography with HiLoad 16/60 Sephadex 200 (Cytiva) in 20 mM HEPES pH 7.5, 300 mM NaCl, 10% glycerol. The His-MBP tag of RBD-CBD variants were removed by incubation with TEV protease overnight in 1:40 mass ratio at 4°C. The untagged proteins were further purified by reverse affinity chromatography with HisPur-Ni-NTA resin in 20 mM HEPES pH 7.5, 300 mM NaCl, 10 mM Imidazole. Lastly, the purified RBD-CBD variants were concentrated and stored in 20 HEPES pH 7.5, 300 mM NaCl, 10% glycerol and 0.5 mM TCEP at -80°C.

### 4.4 Fluorescence conjugation of monoFc-ACE2

Alexa Fluor® 594 conjugation of monoFc-ACE2 was carried out by using Alexa Fluor® 594 Conjugation Kit (Fast) - Lightning-Link® (abcam, ab269822). For each labeling reaction, 100 µL of 1 mg/mL of monoFc-ACE2 in Phosphate Buffer Saline (PBS) pH 7.6 was mixed with 10 µL of Modifier reagent. The 110 µL of mixture was transferred to Alexa Fluor® 594 Conjugation Mix followed by 30 minutes incubation at room temperature in the dark. Then, the reaction was stopped by adding 10 µL of Quencher reagent and for 15 minutes incubation in the dark. Finally, the labelled protein was stored in aliquots of 5 µL at – 80°C freezer before use.

### 4.5 Cellulose pulldown virus neutralization test (cpVNT)

Every testing cassette was assembled by using 1 layer of Whatman No. 1 chromatography paper (GE healthcare, #3001-861) as cellulose test strip and 2 layers of Whatman gel blotting paper, Grade GB005 (GE healthcare, #10426981) as absorbent pads into a cassette housing (Racer Technology Pte. Ltd.). Then, both the test and control spots were blocked with 5 µL of 5% Bovine Serum Albumin (BSA) in PBS pH 7.6. The control spot is further treated with 5 µL of 5 µM RBD-CBD before air-dry. For each test, 20 µL of venous or finger pricked whole blood sample was first incubated with 20 µL of 10 nM RBD-CBD in PBS pH 7.6, 1% BSA for 3 minutes. After that, 40 µL of 5 nM Alexa Fluor594 labelled monoFc-ACE2 (ACE2-AF594) in PBS pH 7.6, 1% BSA was added to the mixture and incubated for another 5 minutes. The final 80 µL reaction was applied equally onto the test and control spot with 40 µL for each. Once sample was fully absorbed, both test and control spots were washed once with 40 µL of PBS pH 7.6. The cassette was then placed in an Atto Testbed for fluorescence measurement. All steps described above were performed at room temperature.

### 4.6 Fluorescence Measurement and Percent Blocking calculation

The Atto Testbed (Attonics Systems Pte Ltd) comprised of an LED lamp (Thorlabs Inc., M590L4), Silicon Avalanche Photodiode detector (SiAPD) (Thorlabs Inc., APD440A) and mCherry filter set (Thorlabs Inc., MDF-MCHA) including an Excitation filter (578/21), an Emission filter (641/75) combined with a dichroic beam-splitter. The testbed was designed specifically to fit the testing cassette dimension for fluorescent signal detection. Fluorescence intensity was recorded as SiAPD output in mV. The percent blocking was calculated using the Equation 1 (see Results section). All samples were tested in triplicates with their mean represented as single data point and the median percent blocking of each group with a given sample size was reported.

### 4.7 Surrogate virus neutralization assay cPass (Genscript)

The assay was performed as per manufacturer’s protocol by first diluting the selected plasma samples 1:10 in the sample dilution buffer provided by the kit, and incubated with HRP-conjugated RBD for 30 minutes at 37°C. Then, the sample-RBD mixtures were transferred to an ACE2 coated ELISA plate for 15 minutes incubation at 37°C before washing with the kit’s washing solution. The sample read-out was performed by adding 100 µL 3,3’,5,5’-tetramethylbenzidine (TMB) solution per reaction well for 15 minutes, followed by 50 µL of stop solution. Absorbance was measured at 450 nm using Infinite 200 PRO multimode TECAN plate reader and the percent of inhibition were calculated according to manufacturer’s recommendation.

### 4.8 Bio-layer Interferometry (BLI)

The streptavidin biosensor tips (Sartoris) were pre-incubated with 20 nM of the monoFc-ACE2, chemically biotinylated with EZ-link Sulfo-NHS-LC-Biotinylation kit (Thermo Fisher, #21435). The binding of WT and all RBD-CBD variants were measured as optical thickness response for 600s of association phase followed by 900s of dissociation phase. The concentration of RBD-CBDs were prepared in serial dilutions ranging from 3.125 - 100 nM (except for gamma RBD-CBD; 2.5 - 80 nM). Analysis of binding response was performed by Octet Data Analysis software using global 1:1 fitting for K_D_ calculation. All experiments were performed using 8-channel Octet RED96e system (Forté Bio) in PBS, 0.2% BSA and 0.05% Tween 20 at 25°C.

### 4.9 SARS-CoV-2 pseudovirus neutralization assay

We applied the same protocol for production of SARS-CoV-2 pseudotyped lentiviral particles and pseudovirus neutralization assay as previously reported ^10^. Briefly, to produce SARS-CoV-2 pseudovirus, HEK293T cells at 36 ×10^6^ cell density were transfected with 27 µg pMDLg/pRRE (Addgene, #12251), 13.5 µg pRSV-Rev (Addgene, #12253), 27 µg pTT5LnX-WHCoV-St19 (SARS-CoV2 Spike) and 54 µg pHIV-Luc-ZsGreen (Addgene, #39196) using Lipofectamine 3000 (Invitrogen, #L3000-150). Then the cells were grown for 3 days in 37º C, 5% CO_2_ incubator. Harvested and filtered viral supernatant were concentrated and quantified by using Lenti-X p24 rapid titer kit (Takara Bio, #632200). Twenty pre and post vaccinated individual plasma were diluted to 1:80 titre with PBS and mixed with equal volume of pseudovirus to 50 µL followed by 1h incubation at 37°C. The neutralization assay was performed by transferring the plasma-pseudovirus mixture to monolayered CHO-ACE2 cells (5×10^4^ cells) in 100µL of complete medium containing DMEM/high glucose with sodium pyruvate (Gibco, #10569010), 10% FBS (Hyclone, # SV301160.03), 10% MEM Non-essential amino acids (Gibco, #1110050), 10% geneticin (Gibco, #10131035) and 10% penicillin/streptomycin (Gibco, #15400054). After 1hr incubation, 150 µL of complete medium were added for subsequent 48hr infection. Each plasma samples were tested in triplicates. The read-out was performed on Tecan Spark 100M after luciferase assay with ONE-glo™ EX reagent (Promega, #E8130) where the percent of neutralization was determined by:

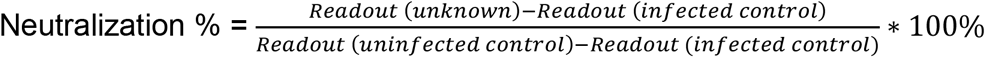

### 4.10 Statistical Analysis

Neutralization antibody (nAb) response was represented by the median % blocking in our results. Since we do not assume a normal distribution, a non-parametric Kruskal-Wallis test with Dunn multiple comparison was performed for comparing Pre-Vac, post 1^st^ dose (1-3 weeks), post 1^st^ dose (3-6 weeks) and post 2^nd^ dose vaccination groups. Meanwhile two-sided Mann-Whitney test was performed for the two-group comparison in analyzing between (i) BNT162b2 and mRNA-1273 post 1^st^ dose (3-6weeks) vaccinated samples and (ii) nAb response within 3 months and post 3months cohorts. The Friedman test with multiple comparison was performed for the same sample set that repeated against RBD-CBD WT and the variants.

## Data Availability

All data produced in the present study are available upon reasonable request to the authors

## ACKNOWLEDGEMENTS

We thank all the IRB-2021-04-020 and DSRB 2012/00917 volunteers for their participation throughout the study. We are grateful for the phlebotomists from National University Hospital, NUS Occupational Health Clinic and Fullerton Healthcare @ NTU for their assistance. The pHLmMBP-10 vector used in the cloning of WT and variant RBD-CBDs was a gift of Luca Jovine, Department of Biosciences and Nutrition, Karolinska Institutet, Sweden. Meanwhile, the TEV protease was a kind gift of NTU Protein Production Platform (proteins.sbs.ntu.edu.sg). We thank Assoc Prof. Tan Yee Joo, Department of Microbiology and Immunology, Yong Loo Lin, School of Medicine, National University of Singapore (NUS) for the CHO-ACE2 stable cell line and SARS-CoV-2 S protein plasmid for the pseudotyped lentiviral particle production. We also greatly appreciate Prof. Herbert Moser and Dr. Felix Neugart from Attonics Inc. for their scientific input. This study is supported by National Health Innovation Singapore (NHIC) grant # NHIC-COVID19-2005004, National Research Foundation via CREATE Share grant #R571-002-021-592 and the Antimicrobial Resistance Interdisciplinary Research Group (AMR-IRG) of Singapore -MIT Alliance in Research and Technology (SMART). All samples acquired from National Centre of Infectious Diseases (NCID) were supported by Singapore Ministry of Health’s National Medical Research Council COVID-19 Research Fund: COVID19RF-0008.

## AUTHOR CONTRIBUTIONS

S.M.L., H.L.C., H.J., P.K., conceived and designed the experimental study. H.J. and P.K. supervised the method development. H.L.C. and S.M.L. performed modified cpVNT experiments and analyzed the data. S.M.L. wrote the manuscript. B. S. performed the pVNT assays. M.W.C. produced the recombinant proteins. S.Y.N prepared the cassettes and cellulose papers. X.G. prepared IRB application and obtained the IRB approval. S.P.T. and S.F.H. designed and developed the portable reader device. J.S., S.S. and B.E.Y. provided the post 3 months vaccinated blood samples. D.M.Y.T edited the manuscript. B.E.Y. M.E.M, H.D.S, P.A.M, P.R.P contributed to the discussion, interpretation of the results and revision of the final manuscript.

## CONFLICT OF INTEREST

PK, MM, HDS, JH and PRP are the cofounders of Thrixen Pte Ltd, a start-up company working in further developing some of the technology presented here.

## FIGURES LEGEND

**Fig. 1: Schematic of cpVNT workflow and results obtained with contrived whole-blood sample. A**. Graphical representation of the modified cpVNT workflow before detection in fluorescent reader. **B**. Cartoon depicting of possible molecular events occur in samples with and without nAb on the test spot and reagent control spot. **C**. Measurement of fluorescence intensity from pre vaccination whole blood samples titrated with 1, 5, 10, 25, 50, 100 nM SARS-CoV-2 monoclonal neutralizing antibody (nAb) **D**. The percent blocking calculated from Equation 1 with nAb concentration presented in log scale.

**Fig. 2: Evaluation of nAb response among pre-vaccination and post-vaccinated individuals using modified cpVNT. A**. The percent blocking measured from 170 whole blood samples of pre vaccination, Pre-Vac (n=36), post 1^st^ dose (1-2W n=10, 3-6W n=50) and post 2^nd^ dose (n=74). The grey line is the median from each group while each dot represents the mean from three independent experiments. **B**. The nAb percent blocking measured in individuals opt for BNT162b2 (n=4 from P1 - 1-2W, n=30 from P1 - 3-6W, n=40 from P2) or **C**. mRNA-1273 (P1 - 1-2W n=6; P1 - 3-6W n=20; P2 n=34). Kruskal-Wallis test with Dunn’s multiple comparison was performed between each vaccination status. **D**. Comparison of nAb percent blocking at P1 3-6W BNT162b2 and P1 3-6W mRNA-1273. Two tailed Mann-Whitney test was performed between the two vaccine brands. The significance values * *P* < 0.05, ** *P* < 0.01, \*\*\**P* < 0.001, \*\*\*\**P* < 0.0001. Total samples: Pre-Vac versus P1 – 3-6W, *P* < 0.001; Pre-Vac versus P2, *P* < 0.0001. BNT162b2: Pre-Vac versus P1 – 3-6W, *P* < 0.05; Pre-Vac versus P2, *P* < 0.0001. mRNA-1273: Pre-Vac versus P1 – 3-6W, *P* < 0.001; Pre-Vac versus P2, *P* < 0.0001. **E**. Twenty-two individual samples percent blocking mapped over pre and post vaccination period comparing between two types of vaccines, BNT162b2 n=12, mRNA 1273 n=10. The window period between first and second dose of vaccination ranged from 4 to 6 weeks depending on individual’s choice.

**Fig. 3: Comparison of modified cpVNT with international standards and established serology tests. A**. The performance of the First WHO International standard Anti-SARS-CoV-2 Immunoglobulin (20/136), Reference Panel for anti-SARS-CoV-2 Mid-tire and Low-titre plasma using modified cpVNT. **B**. The correlation of percent blocking measured from 30 matching plasma and blood samples at pre vaccination (Pre-Vac), post 1^st^ dose (P1) and post 2^nd^ dose (P2) phase using modified cpVNT gave Pearson r, 0.908. The percent blocking for cpVNT with blood samples that correspond to 1000 IU/mL, 220 IU/mL and 44 IU/mL are determined by assuming a linear correlation between the two sample types (see accompanying table). **C**. Comparison of percent blocking measured in the modified cpVNT with percent inhibition of sVNT (cPass™) in 45 matching Pre-Vac, P1 and P2 venous blood and plasma samples. The sensitivity was calculated as 81.5% (CI: 61.9-93.7%), and specificity is 100% (CI: 81.5-100%) when both cpVNT and sVNT’s (cPass™) thresholds were set at 30% blocking. **D**. Comparison between cpVNT and pseudovirus neutralization test (pVNT) with 20 individuals’ sample. The pVNT was performed with plasma in 1:80 dilution. The sensitivity is 100% (CI: 47.8-99.9%) and specificity is 66.7% (CI: 38.4-88.2%) with 30% blocking as the threshold for cpVNT and 50% neutralization for pVNT. All experiments were performed in triplicates.

**Fig. 4: Assessment of nAb response to variants of concern depending on vaccine brand or time post vaccination with our modified cpVNT. A**. Whole blood samples (n=39) from participants who completed vaccination within three months were tested with wildtype (WT) RBD-CBD and variant RBD-CBD. Friedman test with multiple comparison was performed comparing the variants against WT, \**P* < 0.05, \*\**P* < 0.01, \*\*\**P* < 0.001, \*\*\*\**P* < 0.0001. WT versus alpha, *P* < 0.01; WT versus beta, *P* < 0.0001; WT versus gamma, *P* < 0.0001; WT versus delta, n.s. **B**. The percent blocking of nAb measured from 20 P2 samples of BNT162b2 recipients (≤ 3 months) when tested with RBD-CBD variants **C**. Similarly, percent nAb blocking of 19 P2 samples (≤ 3 months) from mRNA-1273 recipients tested with the RBD-CBD variants. **D**. The nAb response in BNT16b2 recipients against WT and RBD-CBD variants when comparing two groups: within three months (n=20) and more than three months (n=37) after complete vaccination. Two-sided Mann Whitney test was performed between the two groups for each variant, WT *P* < 0.0001, alpha *P* < 0.001, beta *P* < 0.0001, gamma *P* < 0.001, delta *P* < 0.0001.

## Notes

### Author Declarations

Institutional Review Board of Nanyang Technological University gave ethical approval for this work (IRB-2021-04-020).

## REFERENCES

1. Bergwerk M, Gonen T, Lustig Y, et al. Covid-19 Breakthrough Infections in Vaccinated Health Care Workers. N Engl J Med. 2021;385:1474–1484. doi:10.1056/NEJMOA2109072

2. Callaway E. COVID vaccine boosters: the most important questions. Nature. 2021;596(7871):178–180. doi:10.1038/d41586-021-02158-6

3. Kamar N, Abravanel F, Marion O, Couat C, Izopet J, Del Bello A. Three Doses of an mRNA Covid-19 Vaccine in Solid-Organ Transplant Recipients. N Engl J Med. 2021;385(7):661–662. doi:10.1056/nejmc2108861

4. Oguntuyo KY, Stevens CS, Hung CT, et al. Quantifying absolute neutralization titers against sars-cov-2 by a standardized virus neutralization assay allows for cross-cohort comparisons of covid-19 sera. MBio. 2021;12(1):1–23. doi:10.1128/mBio.02492-20

5. Gundlapalli A V., Salerno RM, Brooks JT, et al. SARS-CoV-2 serologic assay needs for the next phase of the US COVID-19 pandemic response. Open Forum Infect Dis. 2021;8(1). doi:10.1093/ofid/ofaa555

6. Tan CW, Chia WN, Qin X, et al. A SARS-CoV-2 surrogate virus neutralization test based on antibody-mediated blockage of ACE2–spike protein–protein interaction. Nat Biotechnol. 2020;38(9):1073–1078. doi:10.1038/s41587-020-0631-z

7. Sancilio AE, D’aquila RT, Mcnally EM, et al. A surrogate virus neutralization test to quantify antibody-mediated inhibition of SARS-CoV-2 in finger stick dried blood spot samples. Sci Rep. 123;11:15321. doi:10.1038/s41598-021-94653-z

8. Whitman JD, Hiatt J, Mowery CT, et al. Evaluation of SARS-CoV-2 serology assays reveals a range of test performance. Nat Biotechnol. doi:10.1038/s41587-020-0659-0

9. Wang JJ, Zhang N, Richardson SA, Wu J V. Rapid lateral flow tests for the detection of SARS-CoV-2 neutralizing antibodies. Expert Rev Mol Diagnostic. 2021;21(4):363–370. doi:10.1080/14737159.2021.1913123

10. Kongsuphol P, Jia H, Cheng HL, et al. A rapid simple point-of-care assay for the detection of SARS-CoV-2 neutralizing antibodies. Commun Med. doi:10.1038/s43856-021-00045-9

11. Mattiuzzo G, Bentley EM, Hassall M, et al. Establishment of the WHO International Standard and Reference Panel for anti-SARS-CoV-2 antibody. WHO Expert Comm Biol Stand. Published online 2020. WHO/BS/2020.2403

12. Khoury DS, Cromer D, Reynaldi A, et al. Neutralizing antibody levels are highly predictive of immune protection from symptomatic SARS-CoV-2 infection. Nat Med. 2021;27:1205–1211. doi:10.1038/s41591-021-01377-8

13. Gobeil SMC, Janowska K, McDowell S, et al. Effect of natural mutations of SARS-CoV-2 on spike structure, conformation, and antigenicity. Science (80-). 2021;373(641). doi:10.1126/science.abi6226

14. Laffeber C, de Koning K, Kanaar R, Lebbink JHG. Experimental Evidence for Enhanced Receptor Binding by Rapidly Spreading SARS-CoV-2 Variants. J Mol Biol. 2021;433(15):167058. doi:10.1016/j.jmb.2021.167058

15. Zhu X, Mannar D, Srivastava SS, et al. Cryo-electron microscopy structures of the N501Y SARS-CoV-2 spike protein in complex with ACE2 and 2 potent neutralizing antibodies. PLoS Biol. 2021;19(4):1–17. doi:10.1371/journal.pbio.3001237

16. Wilfredo Garcia-Beltran AF, Lam EC, St Denis K, John Iafrate A, Naranbhai V, Balazs Correspondence AB. Multiple SARS-CoV-2 variants escape neutralization by vaccine-induced humoral immunity. Cell. 2021;184:2372–2383. doi:10.1016/j.cell.2021.03.013

17. Pegu A, O’Connell S, Schmidt SD, et al. Durability of mRNA-1273 vaccine– induced antibodies against SARS-CoV-2 variants. Science. 2021;373(6561):1372–1377. doi:10.1126/science.abj4176

18. Bates TA, Leier HC, Lyski ZL, et al. Neutralization of SARS-CoV-2 variants by convalescent and BNT162b2 vaccinated serum. Nat Commun. 2021;12(5135). doi:10.1038/s41467-021-25479-6

19. Liu J, Liu Y, Xia H, et al. BNT162b2-elicited neutralization of B.1.617 and other SARS-CoV-2 variants. Nature. 2021;596(7871):273–275. doi:10.1038/s41586-021-03693-y

20. Pouwels KB, Pritchard E, Matthews P, et al. Effect of Delta on viral burden and vaccine effectiveness against new SARS-CoV-2 infections in the UK. Nat Med. Published online 2021. doi:10.1038/s41591-021-01548-7

21. Harvey WT, Carabelli AM, Jackson B, et al. SARS-CoV-2 variants, spike mutations and immune escape. Nat Rev Microbiol. 2021;19:409–424. doi:10.1038/s41579-021-00573-0

22. Liu C, Ginn HM, Dejnirattisai W, et al. Reduced neutralization of SARS-CoV-2 B.1.617 by vaccine and convalescent serum. Cell. 2021;184(16):4220-4236.e13. doi:10.1016/j.cell.2021.06.020

23. Greaney AJ, Starr TN, Barnes CO, et al. Mapping mutations to the SARS-CoV-2 RBD that escape binding by different classes of antibodies. Nat Commun. 2021;12(1). doi:10.1038/s41467-021-24435-8

24. Tan AT, Lim JME, Bert N Le, et al. Rapid determination of the wide dynamic range of SARS-CoV-2 Spike T cell responses in whole blood of vaccinated and naturally infected. J Clin Invest. 2021;131(17). doi:10.1172/JCI152379.

25. Piccoli L, Park YJ, Tortorici MA, et al. Mapping Neutralizing and Immunodominant Sites on the SARS-CoV-2 Spike Receptor-Binding Domain by Structure-Guided High-Resolution Serology. Cell. 2020;183(4):1024-1042.e21. doi:10.1016/j.cell.2020.09.037

26. Robbiani DF, Gaebler C, Muecksch F, et al. Convergent antibody responses to SARS-CoV-2 in convalescent individuals. Nature. 2020;584(7821):437–442. doi:10.1038/s41586-020-2456-9

27. Shi R, Shan C, Duan X, et al. A human neutralizing antibody targets the receptor-binding site of SARS-CoV-2. Nature. 2020;584(7819):120–124. doi:10.1038/s41586-020-2381-y

28. Uriu, Keiya; Kimura, Azumi; Shirakawa, Kotaro; Takaori-Kondo, Akifumi; Nakada, Taka-aki; Kaneda, Atsushi; Nakagawa, So; Sato K. Neutralization of the SARS-CoV-2 Mu variant by convalescent and Vaccine Serum. N Engl J Med. Published online 2021. doi:10.1056/NEJMc2114706

29. Mizrahi B, Lotan R, Kalkstein N, et al. Correlation of SARS-CoV-2 Breakthrough Infections to Time-from-vaccine. Nat Commun. 2021;12:6379. doi:10.1038/s41467-021-26672-3

