## Supplemental Figures for "Finger stick blood test to assess post vaccination SARS-CoV-2 neutralizing antibody response against variants"

### Supporting Information

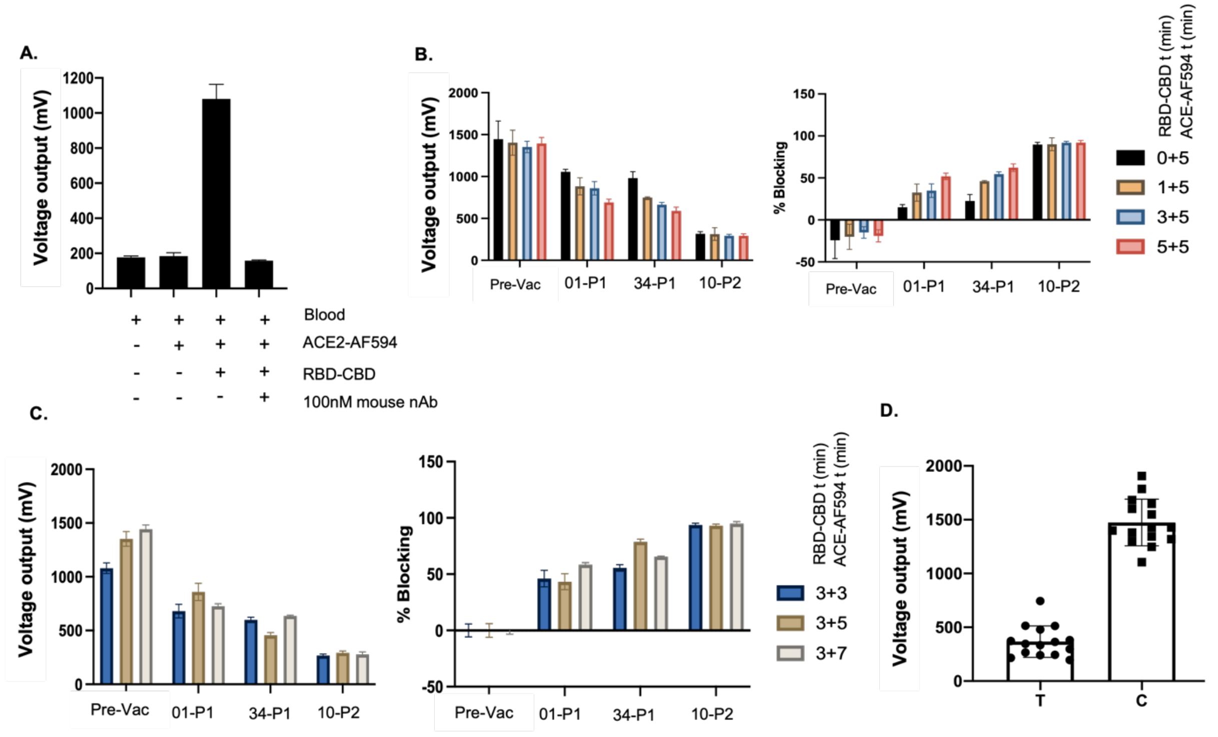


**Fig. S1. Optimization of modified cpVNT with Alexa Fluor594 labeled ACE2.**
**A.** Fluorescence intensity (voltage output) measured with blood samples in the presence and absence of mouse monoclonal SARS-CoV-2 nAb. **B.** Four different participants' samples: pre-vaccination (Pre-Vac), post 1$^{st}$ dose (01-P1, 34-P1) and post 2$^{nd}$ dose (P2) were used for optimization of RBD-CBD incubation time and **C.** ACE2-AF594 incubation time. All samples were tested in triplicates. **D**. Comparison of fluorescent intensity between Test spot-T and Control spot-C with 15 post 2nd dose vaccinees' whole blood samples.

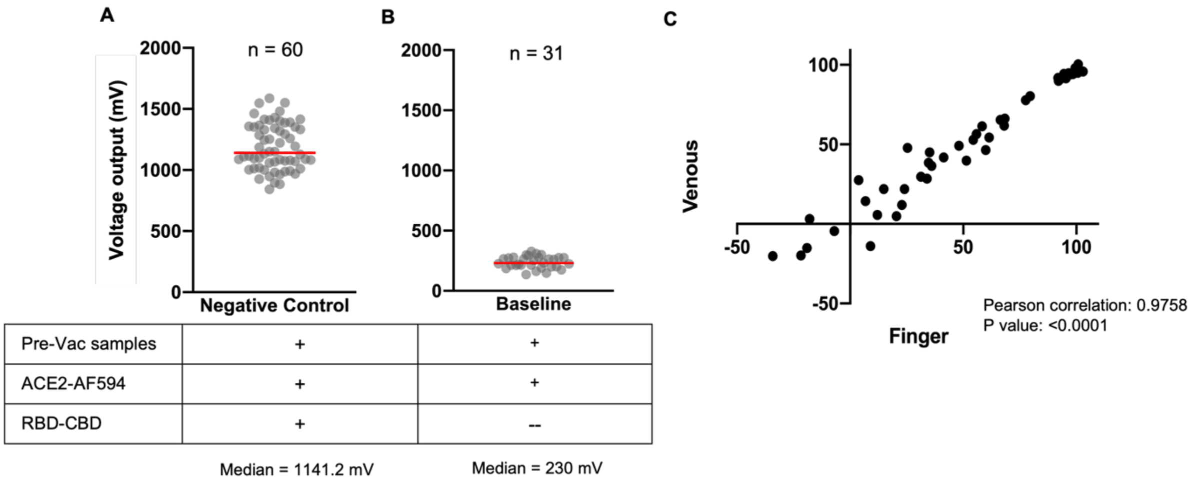


**Fig. 2. Signal intensity for negative control and baseline as well as correlation of plasma and whole blood samples with modified cpVNT.**
**A**. Fluorescence intensity (voltage output) measured from 60 pre-vaccination blood samples, red line represents median of the distribution, 1141 mV; which is the value used as NC for % blocking calculation. **B**. Total 31 samples (pre and post vaccination) were measured in the absence of RBD-CBD, where the red line is the median, 230 mV. **C.** A total of 46 matching samples of finger stick and venous blood showed strong correlation (Pearson r: 0.9758, p< 0.0001) in percent blocking detection using the modified cpVNT.

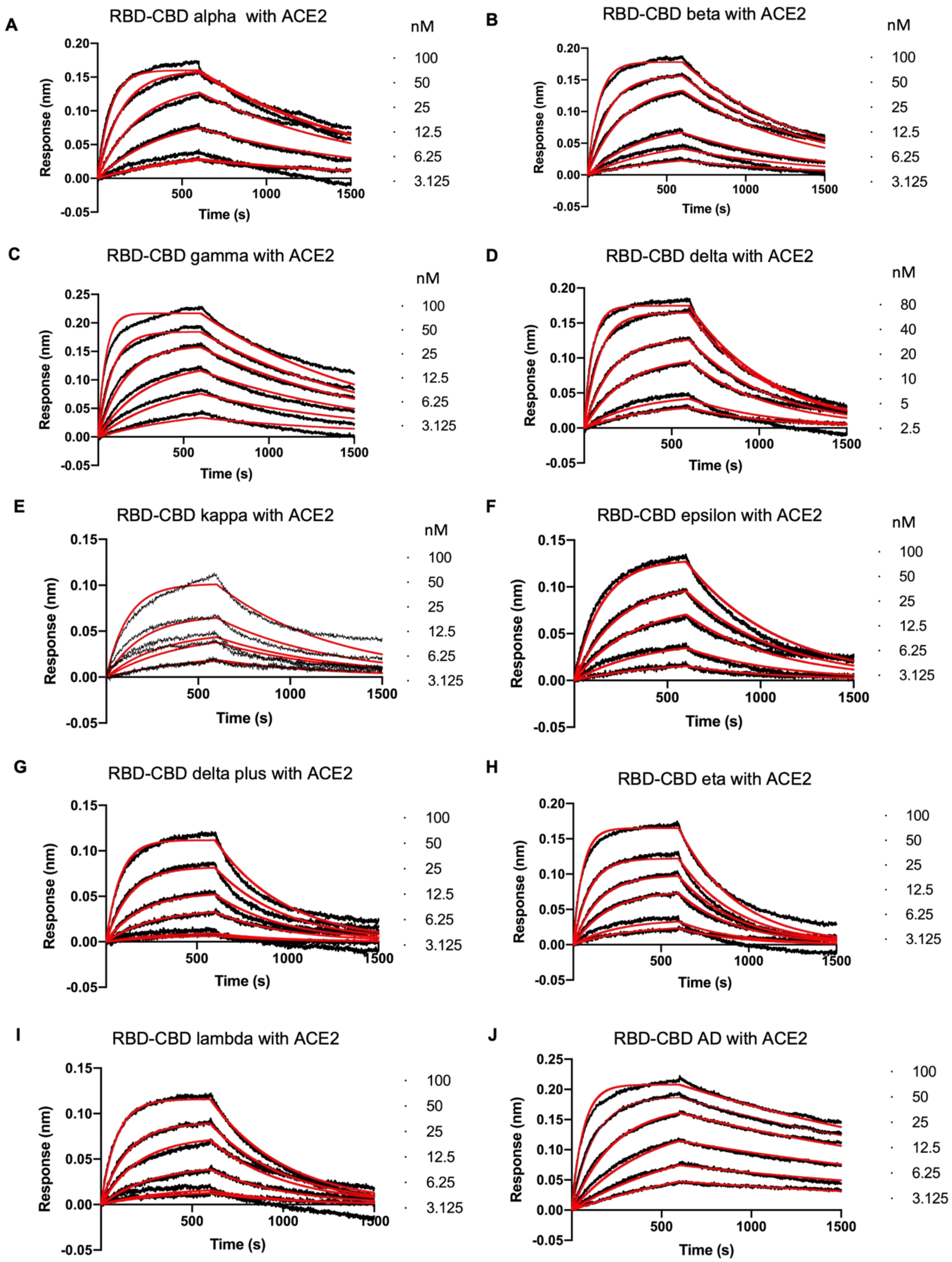


**Fig. S3 Biolayer interferometry profiles for RBD-CBD variants with biotinylated ACE2 immobilized on streptavidin biosensors.**

**A.** RBD-CBD alpha, **B**. RBD-CBD beta, **C**. RBD-CBD gamma, **D**. RBD-CBD delta, **E**. RBD-CBD kappa, **F.** RBD-CBD epsilon, **G.** RBD-CBD delta plus, **H.** RBD-CBD eta, **I.** RBD-CBD lambda and **J.** RBD-CBD AD

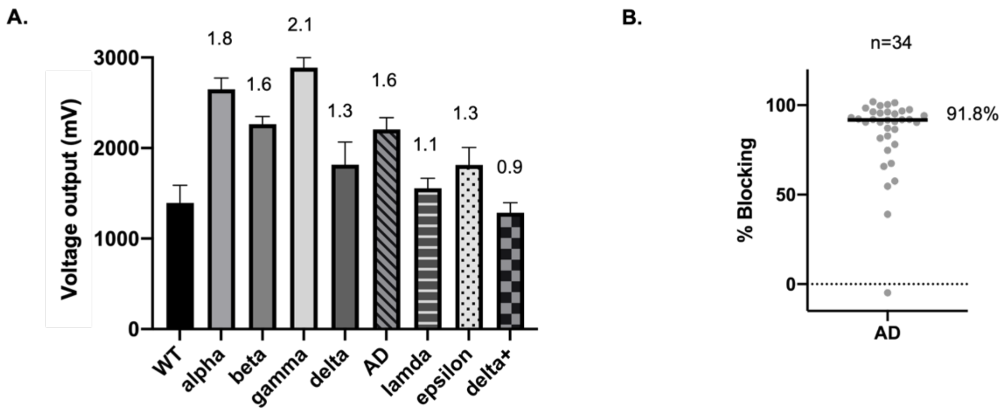


**Fig. S4 Signal intensity of RBD-CBD variants' response with fluorescence labelled ACE2.**

**A**. The fluorescence intensity (voltage output) of pre vaccination blood sample when tested against wildtype (WT) and RBD-CBD variants of different binding affinity against ACE2. The numbers above each bar indicate fold of increase in intensity as compared to WT. **B.** The nAb percent blocking measured from 34 post 2$^{nd}$ dose vaccinated samples (within 3 months) against AD, the hypothetical RBD-CBD.

**Table S1. Fluorescence intensity (voltage output) measured from two batches of ACE2-AF594 with pre vaccinate blood samples for negative control calibration.**

| ACE2-AF594 Batch | RBD-CBD Concentration (nM) | Fluorescence intensity for NC | # Samples | Fluorescence intensity for baseline | # Samples |
|---|---|---|---|---|---|
| 1 | 10 | 1088.7 (±179.8) | 17 | 181.1 (±18.3) | 7 |
| 2 | 10 | 1235.5 (±185.9) | 43 | 254.1 (±25.9) | 24 |

**Table S2: Samples used for modified cpVNT-blood in comparison with pVNT, cPass and cpVNT-plasma**

| Sample ID | Vaccination status | % Neutralization in pVNT | % Inhibition in cPass | % Blocking cpVNT-blood | % Blocking cpVNT-plasma |
|---|---|---|---|---|---|
| 0001-V-V05-PV | Pre-Vac | | -7.763312 | 22.94239 | |
| 0003-F-V03-PV | Pre-Vac | | -17.49623 | -18.20988 | |
| 0004-V-V01-PV | Pre-Vac | 0 | | 19.75356 | 19.01346 |
| 0004-V-V04-PV | Pre-Vac | | -2.99429 | -7.40E-15 | |
| 0004-F-V06-PV | Pre-Vac | | -5.805046 | 12.53561 | |
| 0005-V-V01-PV | Pre-Vac | | -13.7654 | 27.02332 | |
| 0007-V-V01-PV | Pre-Vac | 0 | -10.4598 | 11.35117 | 11.89958 |
| 0008-V-V01-PV | Pre-Vac | 0 | -14.8892 | 11.96845 | -21.78846 |
| 0009-V-V01-PV | Pre-Vac | | -8.86967 | 13.20597 | |
| 0011-V-V01-PV | Pre-Vac | | -11.27776 | -27.88066 | |
| 0013-F-V06-PV | Pre-Vac | | -5.521211 | -12.65432 | |
| 0013-F-V08-PV | Pre-Vac | 0 | | 8.40192 | |
| 0022-F-V01-PV | Pre-Vac | | 1.500837 | -11.34092 | |
| 0022-V-V01-PV | Pre-Vac | 0.12 | 11.52455 | 21.31724 | 15.95626 |
| 0026-F-V01-PV | Pre-Vac | | -6.576852 | -38.8203 | |
| 0027-F-V01-PV | Pre-Vac | | 3.549902 | 12.41427 | |
| 0030-F-V01-PV | Pre-Vac | | 7.101754 | 14.3 | |
| 0036-V-V01-PV | Pre-Vac | 0 | | 9.84225 | 11.50885 |
| 0042-V-V01-PV | Pre-Vac | | | 12.89438 | 8.254454 |
| 0054V-V01-PV | Pre-Vac | | | 1.646091 | -4.238932 |
| 0001-F-V09-P1-02W-MD | P1 | | 66.0465 | 31.893 | |
| 0001-V-V12-P1-05W-MD | P1 | 43.56 | 72.68333 | 36.07682 | 73.514022 |
| 0003-V-V06-P1-03W-PF | P1 | 1.03 | 44.83542 | 21.46776 | 17.1909 |
| 0004-V-V10-P1-03W-MD | P1 | | 75.51875 | 22.39369 | |
| 0009-V-V02-P1-03W-PF | P1 | 3.21 | 10.31425 | 28.45651 | 8.348457 |

| | | | | | |
|---|---|---|---|---|---|
| 0010-V-V03-P1-01W-MD | P1 | | -8.83867 | 22.97668 | 19.5426 |
| 0010-F-V06-P1-03W-MD | P1 | | 89.66134 | 73.07956 | |
| 0011-V-V02-P1-03W-PF | P1 | | 31.8281 | 34.12209 | |
| 0012-V-V01-P1-03W-PF | P1 | 3.02 | 41.51574 | 39.16324 | 28.42025 |
| 0013-F-V09-P1-02W-MD | P1 | | 67.62917 | 20.78189 | |
| 0019-V-V2-P1-03W-PF | P1 | 37.75 | 63.88273 | 27.02332 | 50.4968 |
| 0020-F-V01-P1-04W-PF | P1 | | 81.96858 | 31.48148 | |
| 0022-F-V02-P1-01W-PF | P1 | | 1.500837 | 10.90535 | |
| 0022-V-V04-P1-06W-PF | P1 | 37.75 | 71.37292 | 41.46091 | 60.19754 |
| 0025V-V03-P1-6W-PF | P1 | | | 48.76543 | 53.05427 |
| 0045V-V05-P1-5W-PF | P1 | | | 61.21399 | 64.01905 |
| 0046V-V01-P1-3W-MD | P1 | | | 80.55843 | 96.97219 |
| 0049-V-V01-P1-03W-MD | P1 | | 79.53542 | 39.47188 | 52.93668 |
| 0052-V-V01-P1-05W-PF | P1 | 36.48 | 72.69792 | 28.01783 | 54.58287 |
| 0053V-V01-P1-5W-PF | P1 | 44.46 | 71.42917 | 38.13443 | 74.68987 |
| 0054-V-V02-P1-03W-PF | P1 | | | 43.34705 | 33.47639 |
| 0062V-V01-P1-5W-MD | P1 | | | 75.5144 | 83.94967 |
| 0001-V-V16-P2-02W-MD | P2 | | 96.88125 | 95.81619 | 99.88242 |
| 0002-V-V07-P2-07W-PF | P2 | | 97.30353 | 104.3553 | |
| 0002-V-V09-P2-14W-PF | P2 | 96.19 | 96.91042 | 90.84362 | 99.58845 |
| 0004-F-V12-P2-01W-MD | P2 | | 71.09583 | 40.9808 | |
| 0004-V-V14-P2-02W-MD | P2 | 97.46 | 96.775 | 91.80384 | 99.94121 |
| 0006-V-V01-P2-01W-MD | P2 | 48.63 | 91.3985 | 90.18098 | |

| | | | | | |
|---|---|---|---|---|---|
| 0007-V-V08-P2-03W-MD | P2 | 94.56 | 96.81875 | 91.02304 | 95.18118 |
| 0009-V-V03-P2-03W-PF | P2 | 63.7 | 93.34792 | 84.22497 | |
| 0009-V-V04-P2-12W-PF | P2 | | | 56.15305 | 63.6075 |
| 0010-F-V07-P2-01W-MD | P2 | | 94.50406 | 88.40878 | |
| 0024-F-V01-P2-08W-PF | P2 | | 96.54536 | 85.69959 | |
| 0028-F-V01-P2-09W-PF | P2 | | 92.88269 | 81.44719 | |
| 0029-F-V01-P2-05W-PF | P2 | | 81.82083 | 63.3059 | |
| 0034-V-V03-P2-02W-MD | P2 | 99.64 | 97.06042 | 96.46776 | 99.88242 |
| 0046V-V02-P2-3W-MD | P2 | | | 94.20439 | 95.70815 |
| 0067V-V01-P2-3W-PF | P2 | | | 79.13328 | 61.02064 |
| **Total samples** | | **20** | **45** | **58** | **30** |

**Table S3: The percent inhibition of WHO International Standard and Reference Panel's plasma measured using cPass™ .**

| WHO Plasma Standard & References | Neutralizing activity (IU/mL) | % Inhibition measured in cPass |
|---|---|---|
| International Standard | 1000 | 94.3 (±0.26) |
| International Standard (1:1 dilution) | 500 | 90.6 (±0.73) |
| Mid titre | 210 | 78.7 (±0.65) |
| Low titre | 44 | 19.0 (±3.2) |
| Low titre (1:1 dilution) | 22 | 8.3 (±1.4) |
